# Improving research transparency with individualized report cards: A feasibility study in clinical trials at a large university medical center

**DOI:** 10.1101/2024.02.10.24302619

**Authors:** Delwen L. Franzen, Maia Salholz-Hillel, Stephanie Müller-Ohlraun, Daniel Strech

**Author notes:** These authors contributed equally to this work (Delwen L. Franzen, Maia Salholz-Hillel). Author order was determined alphabetically.

## Abstract

Research transparency is crucial for ensuring the relevance, integrity, and reliability of scientific findings. However, previous work indicates room for improvement across transparency practices. The primary objective of this study was to develop an extensible tool to provide individualized feedback and guidance for improved transparency across phases of a study. Our secondary objective was to assess the feasibility of implementing this tool to improve transparency in clinical trials.

We developed study-level “report cards” that combine tailored feedback and guidance to investigators across several transparency practices, including prospective registration, availability of summary results, and open access publication. The report cards were generated through an automated pipeline for scalability. We also developed an infosheet to summarize relevant laws, guidelines, and resources relating to transparency. To assess the feasibility of using these tools to improve transparency, we conducted a single-arm intervention study at Berlin’s university medical center, the Charité – Universitätsmedizin Berlin. Investigators (n = 92) of 155 clinical trials were sent individualized report cards and the infosheet, and surveyed to assess their perceived usefulness. We also evaluated included trials for improvements in transparency following the intervention.

Survey responses indicated general appreciation for the report cards and infosheet, with a majority of participants finding them helpful to build awareness of the transparency of their trial and transparency requirements. However, improvement on transparency practices was minimal and largely limited to linking publications in registries. Investigators also commented on various challenges associated with implementing transparency, including a lack of clarity around best practices and institutional hurdles.

This study demonstrates the potential of developing and using tools, such as report cards, to provide individualized feedback at scale to investigators on the transparency of their study. While these tools were positively received by investigators, the limited improvement in transparency practices suggests that awareness alone is likely not sufficient to drive improvement. Future research and implementation efforts may adapt the tools to further practices or research areas, and explore integrated approaches that combine the report cards with incentives and institutional support to effectively strengthen transparency in research.

## Introduction

The inability to reproduce scientific findings (Baker, 2016; Begley and Ellis, 2012; National Academies of Sciences, Engineering, and Medicine, 2019; Open Science Collaboration, 2015) and the non-or incomplete reporting of research results (Bassler et al., 2016; Franco et al., 2014; Ioannidis et al., 2014; Song et al., 2010; Toews et al., 2017) have raised concerns about research waste, and highlighted the need to promote responsible research practices as a means to increase research value. Transparency is core to responsible research: transparency practices allow the quality of research to be appraised, mitigate biases, discourage poor methodological practices, and facilitate comprehensive evidence synthesis (Bradley et al., 2020; Glasziou et al., 2014; Munafò et al., 2017; Nosek et al., 2015). The relevance of these practices is emphasized in initiatives to reform research assessment (Hicks et al., 2015; “San Francisco Declaration on Research Assessment (DORA),” 2012), which have included calls to increase recognition of researchers who commit to robust and transparent practices (Moher et al., 2020). One of the principles underlying the 2022 Agreement on Reforming Research Assessment is to focus research assessment criteria on quality, where “research is carried out through transparent research processes and methodologies” (Coalition for Advancing Research Assessment, 2022).

Reforming how research is performed and assessed requires evaluation and communication about the current uptake of responsible research practices, including transparency. Such monitoring efforts largely focus on a single phase of the research process (e.g., results reporting) and communicate findings at a group level, such as institution or country. For example, the EU Trials Tracker shows compliance of clinical trials with results reporting requirements and presents these data as a dashboard at the level of institutional sponsors (Goldacre et al., 2018). In turn, the French Open Science Monitor shows the evolution at the national level of several open science practices, such as open access and mentions of data sharing in publications (Jeangirard, 2019). These initiatives and others (Barbers et al., 2022; Diprose et al., 2023; Franzen et al., 2023; QUEST Center for Responsible Research, Berlin Institute of Health at Charité Universitätsmedizin Berlin, n.d.) provide baseline information against which change can be measured and empower efforts to drive research transparency.

Policy and monitoring activities reflect top-down approaches to inform and trigger change in research transparency. Yet improvement ultimately requires researchers to take specific actions, such as report results and methods in a timely manner and in sufficient detail. Thus, effective research reform also requires bottom-up approaches to empower researchers through feedback and guidance on transparency at the level of individual studies. Efforts to increase the transparency of individual studies largely focus on publications. Journals have experimented with awarding authors open science badges to incentivize practices such as open data (Kidwell et al., 2016; Rowhani-Farid et al., 2020). The ScreenIT pipeline screens preprints in medRvix and bioRxiv for common problems in how research is reported and provides rapid feedback to help authors improve transparency and reproducibility (Schulz et al., 2022).

Building on these efforts, we aimed for a complementary approach to provide feedback and guidance to investigators on transparency practices associated with different phases of a study. While such a feedback tool would be beneficial across research areas and study types, our feasibility study focused on clinical trials. Clinical trials are well suited to evaluate the feasibility and usefulness of such a tool for both normative and technical reasons. The significance of clinical trials to patient health has inspired ethical guidelines and legal requirements for transparency practices across the research lifecycle, from prospective registration to timely results in registries and journal articles (International Committee of Medical Journal Editors, 2023; World Health Organization, 2017a; World Medical Association, 2013). In turn, clinical trial registries provide a ready list of studies and collect data relating to those trials, from study start to completion, which enables evaluation of these practices (Saberwal, 2021; World Health Organization, 2017b; Zarin et al., 2017). In previous work, we developed tools to monitor and communicate performance on these practices at the level of institutions, which could be adapted to the level of individual studies (Franzen et al., 2023).

This study’s primary objective was to develop a scalable tool to provide feedback and guidance on transparency across research phases of an individual study. We designed individualized “report cards” that summarize the performance of a given clinical trial across several transparency practices and provide tailored guidance for improvement. We also created an infosheet with an overview of recommended transparency practices and relevant guidelines and regulations.

The secondary objective was to investigate the feasibility of implementing these tools to improve trial transparency. We conducted an exploratory, single-arm intervention and disseminated individualized trial transparency report cards and the infosheet to investigators at the Charité – Universitätsmedizin Berlin, in collaboration with the institutional Clinical Study Center. We surveyed investigators on the self-reported usefulness of the materials and evaluated the included trials for improvements in transparency.

## Results

### Tools: report cards and infosheet

We developed study-level report cards that combine individualized feedback and guidance on the transparency of a given trial (Figure 1 (a)). The report cards provide feedback on the following transparency practices: a) prospective registration, b) availability and timeliness of summary results in the registry, c) availability and timeliness of a results publication (earliest if multiple), and if a publication exists, d) links between registration and the results publication, and e) open access status of the publication. In case of a validated cross-registration in the EU Clinical Trials Register (EUCTR), the report card additionally displays the following practices based on the EUCTR registration: a) prospective registration, and b) availability of summary results in the registry. For transparency practices that can still be improved after trial completion, the report card includes a call-to-action and a link to resources to implement the practice (e.g., instructions in the relevant registry).

**Figure 1:**
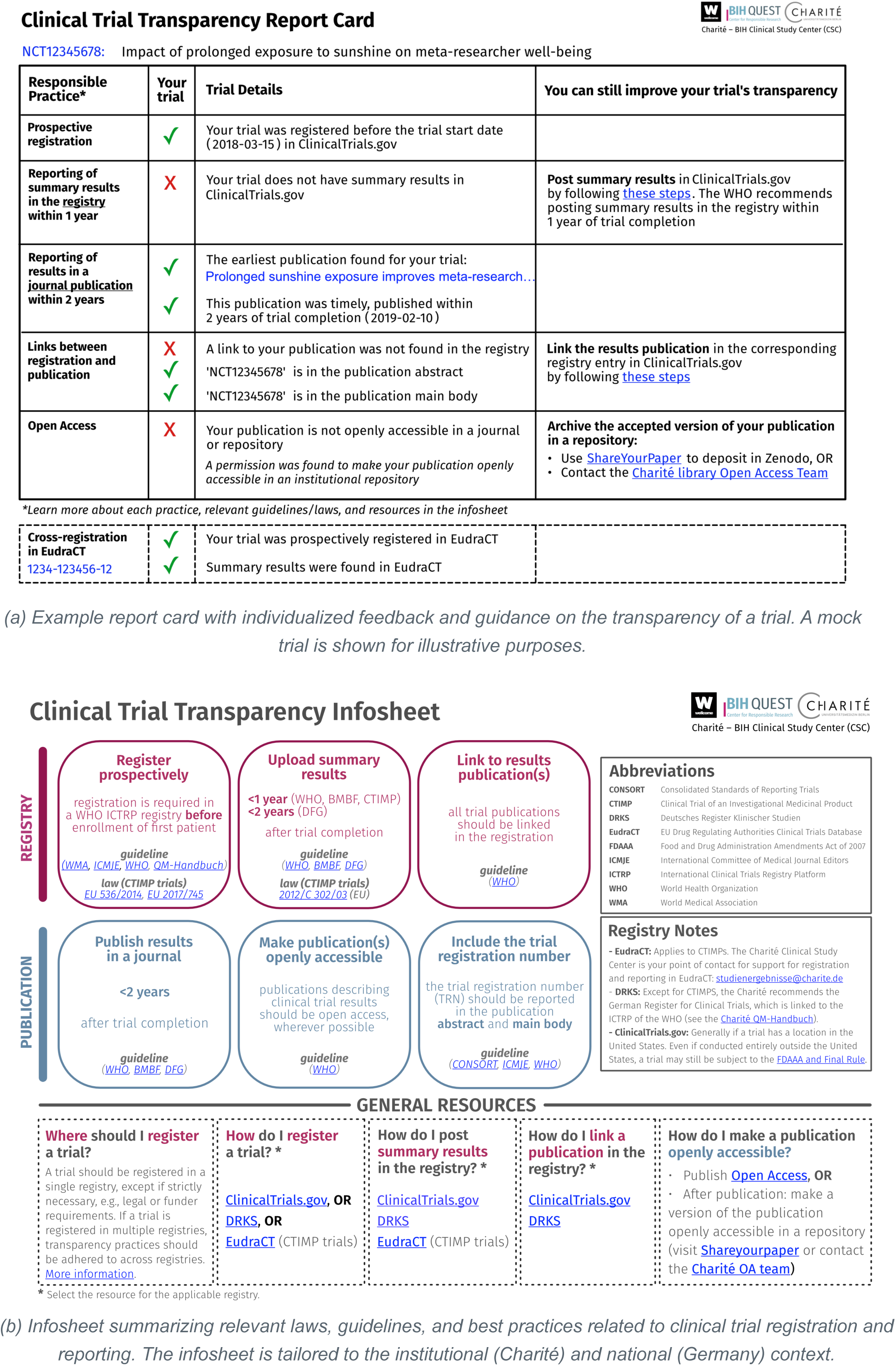
Clinical trial transparency tools including (a) report card and (b) infosheet.

We also developed an infosheet with an overview of guidelines and laws relating to clinical trial transparency, tailored to the national and institutional contexts (Figure 1 (b)). The infosheet summarizes transparency practices relating to trial registration and reporting, their normative basis, and available resources to implement each practice.

The final versions of the report card and infosheet incorporated feedback from expert reviews and cognitive interviews. Suggestions for improvement from participants largely related to the visual design, scope, and implementation of the tools. Themes are elaborated in Supplement 1 Table 1.

### Feasibility study

#### Sample characteristics

To investigate the feasibility of using report cards and an infosheet to improve trial transparency, we started with a dataset of 2,909 trials generated in previous studies (known as “IntoValue”; see details in the Methods). After limiting this set to trials led by the Charité and completed between 2014 and 2017, we contacted 128 investigators responsible for 168 trials. After excluding unreachable investigators, our analysis sample included 92 investigators responsible for 155 trials. Figure 2 (a) details screening of both trials and investigators. Characteristics of the included trials and investigators are in Supplement 2 Table 1.

**Figure 2:**
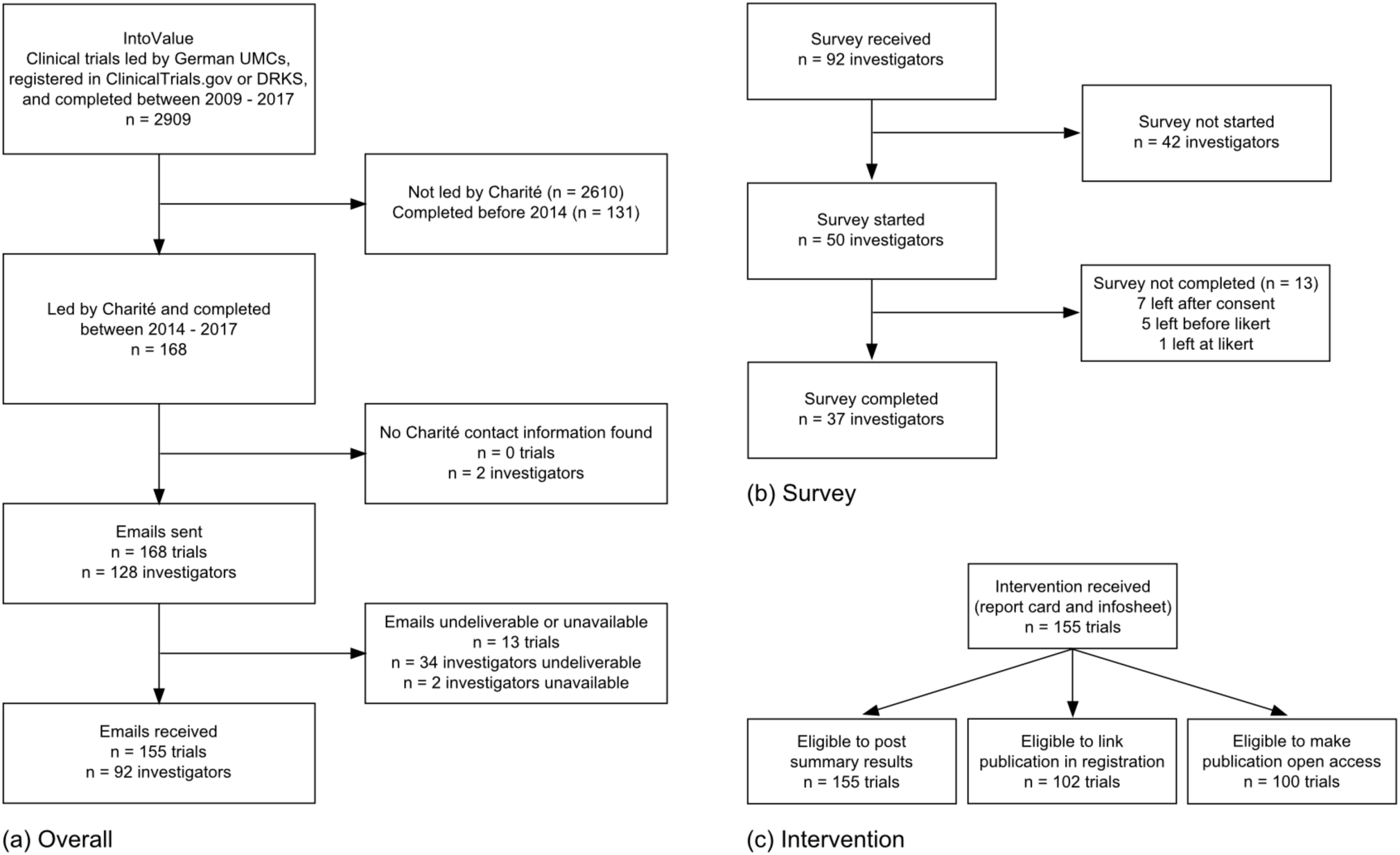
Screening flowchart of trials and trial investigators. (a) shows overall screening. (b) shows screening of investigators for the survey analysis. (c) shows screening of trials for the analysis of improvement in transparency practices; eligible trials include trials that had already completed the practice at intervention launch.

#### Survey

A total of 37 researchers (40% of the 92 contacted) completed the survey in the 12 weeks between survey launch on 25 May 2022 and study close on 17 August 2022. Figure 2 (b) shows a flowchart of investigators and exclusion from analysis at each stage. Supplement 3 Figure 1 shows the response timeline across the survey fielding period. The survey took respondents generally just under 4 minutes, with a median response time of 237 seconds (IQR: 124 - 402). The majority of respondents self-reported as study leads or study doctors (n = 32, 86%); see Supplement 3 Figure 2 for additional details on respondent The survey asked trial investigators to rate their agreement with each of eight statements about the report card and infosheet (for example, “The infosheet is clear.”). Respondents shared overall positive perceptions about the report card and infosheet, with at least 49% agreeing or strongly agreeing with all provided statements, and mean scores on all individual items ranging from 3.2 to 3.7, on a scale from 1 to 5. A peak of 73% of respondents found the infosheet with an overview of relevant laws, guidelines, and resources generally valuable. The statement with the lowest agreement (49%) and highest disagreement (38%) was whether the report cards were helpful to improve their own trial’s transparency. Figure 3 shows responses to each item; means with 95% confidence intervals are presented in Supplement 3 Figure 3.

**Figure 3:**
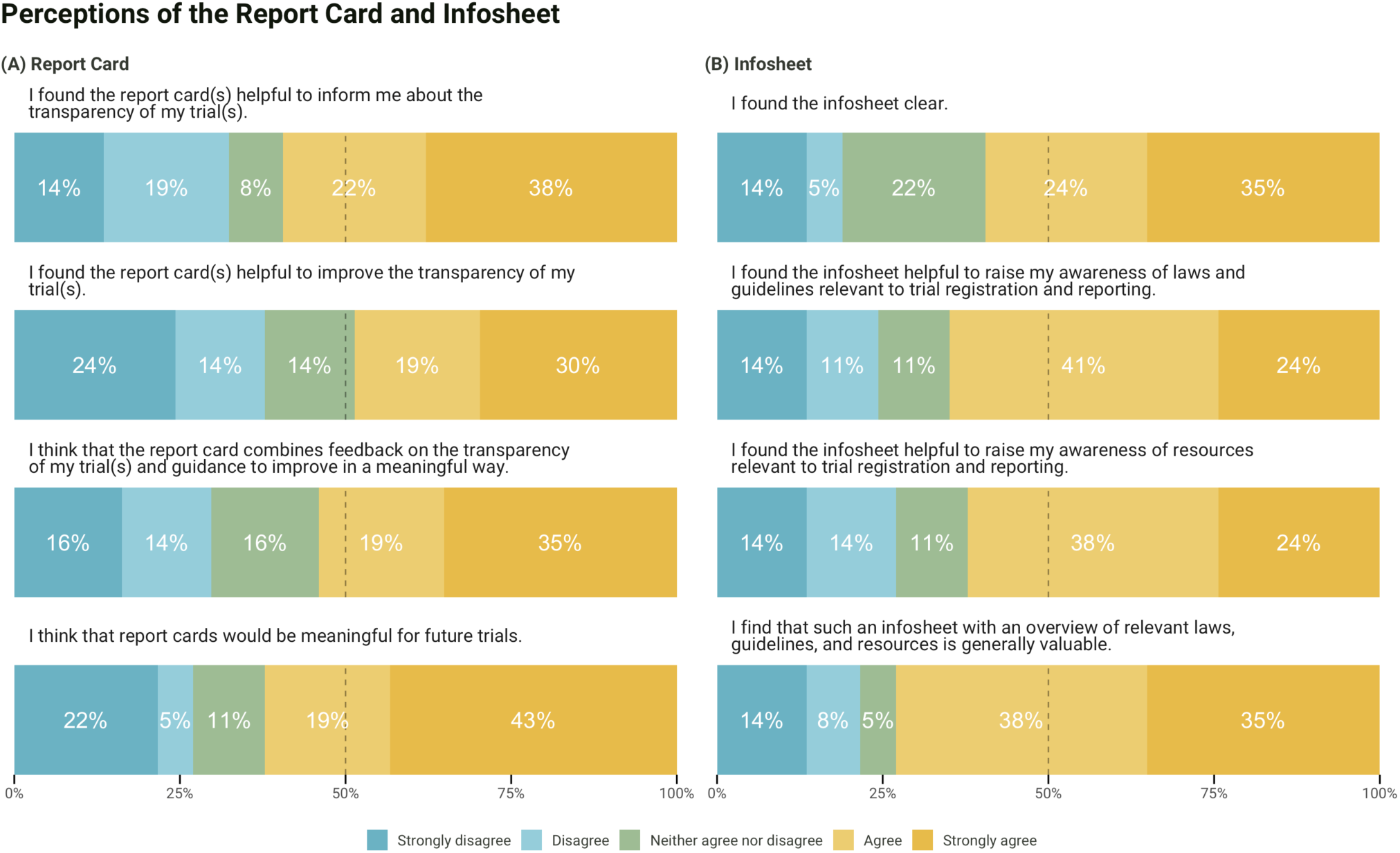
Perceptions of the Report Card and Infosheet.

A subgroup analysis of 3 participants who reported not reviewing the materials before starting the survey showed the most negative option (i.e., strongly disagree) across all statements on both the report card and infosheet. Another subgroup analysis showed no meaningful difference between study leads and doctors (n = 32) versus other respondents (n = 5). See Supplement 3 for further details on these subgroup analyses.

Respondents were invited to share comments on the materials. After removing non-substantive responses (e.g., “I have no comment”), 17 remained. Comments included positive remarks and suggestions for improvement of the materials, as well as broader challenges around the management and coordination of clinical trials. Supplement 3 Table 1 details the suggestions and challenges provided in comments.

Investigators also had the opportunity to provide corrections to their report card. In our manual review of the 18 provided corrections, we found that only 5 were valid, while the remaining were incorrect understandings of the practices or additional information on the trials beyond the scope of the report card. See Supplement 3 for details.

#### Improvement in transparency practices

Trial transparency improved minimally across the follow-up period for eligible trials (see Figure 2 (c) for a screening flowchart for eligible trials). The largest improvement was in linked publications, which increased 5.9%: 5 trials added links to their publications in the registry in the 3 months following our intervention, and 1 additional trial did so within 1 year of the intervention. One of these trials also posted summary results in the registry. While these changes were small, no changes were seen in the 3-months preceding the intervention (Figure 4). Additionally, one publication became openly accessible on the journal website and not a repository, suggesting this was not triggered by an investigator and was hence independent of our intervention.

**Figure 4:**
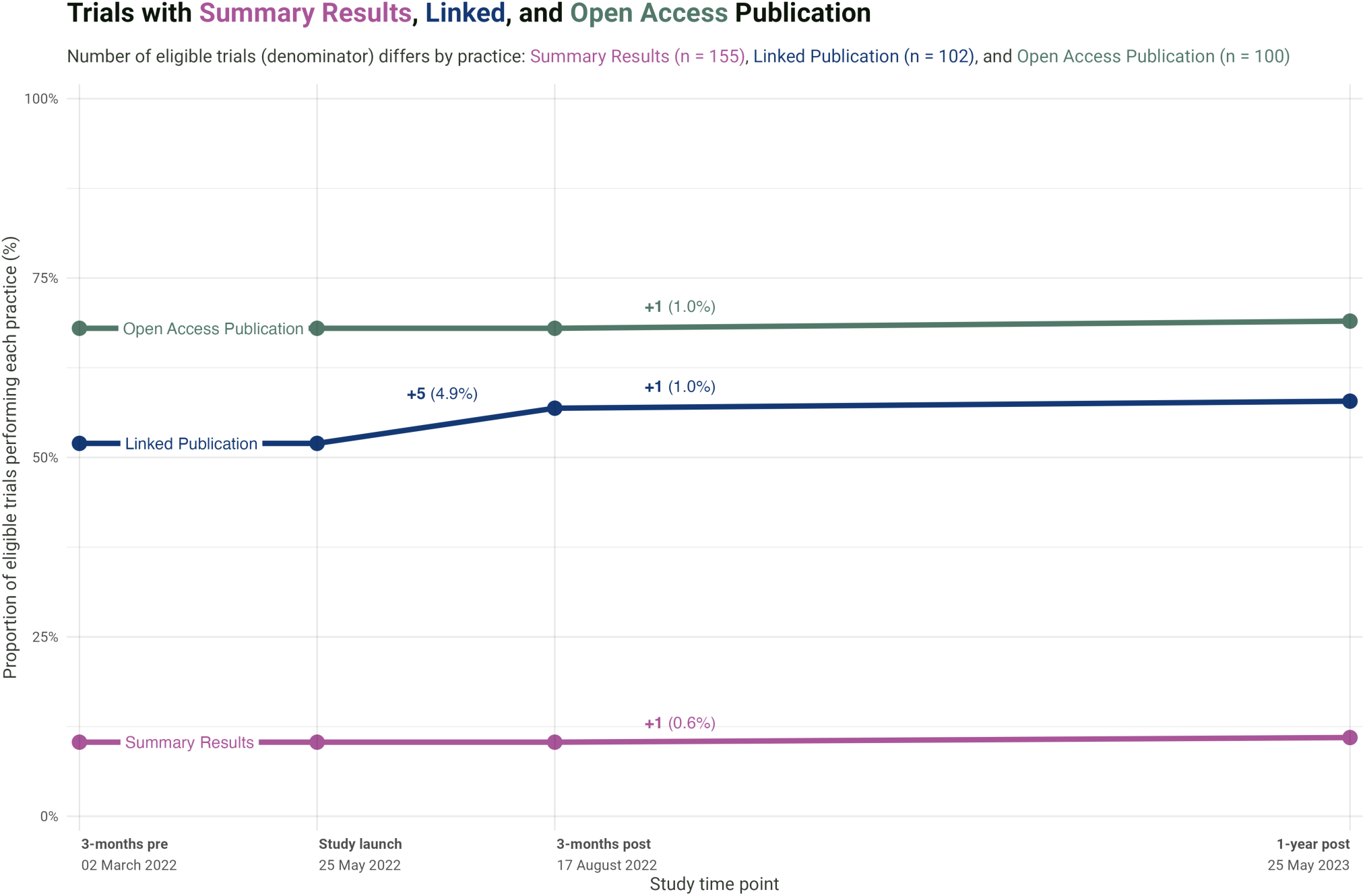
Overall proportion of trials performing each practice at each study time point (3 months prior to study launch, at study launch, and after study launch at 3 and 12 months). The different denominators across practices reflect distinct requirements to perform each practice. Summary results: denominator is all trials (n = 155). Linked publication: denominator is all trials with a publication (n = 102). Open access publication: denominator is all trials with a publication and a publisher permission to self-archive the paper in an institutional repository (n = 100).

## Methods

### Tools: report cards and infosheet

#### Design

We set out to develop report cards with individualized feedback on the transparency of a given trial. The report cards aimed to: a) provide feedback on registration and reporting best practices at the level of an individual trial, b) indicate visually (e.g., tick mark, cross) whether a practice was followed, c) include the trial details relevant to each practice, d) be succinct (max. 1 page), and e) be generated with open source code for scalability. We also designed an infosheet that provides an overview of guidelines and laws relating to clinical trial transparency. The infosheet aimed to: a) provide an overview of the recommended or required transparency practices and their normative basis (i.e., guidelines or laws), b) include links to resources for each practice, c) be tailored to the German and institutional context, and d) be succinct (max. 1 page). The design of the report cards and infosheet was shaped by feedback obtained in cognitive interviews and from experts in clinical trial transparency Supplement 1. The report cards and infosheet were presented in English.

#### Trial transparency dataset

The report cards require a structured trial transparency dataset, i.e., a dataset of clinical trials and their performance across several transparency practices. We previously developed methods to assess transparency practices for a cohort of trials using a combination of automated and manual approaches, which are described in detail elsewhere (Franzen et al., 2023; Franzen, 2023; Riedel et al., 2022; Salholz-Hillel et al., 2022). Supplement 4 summarizes the main steps to prepare a trial transparency dataset for use with the report cards.

#### Technical implementation: translating the dataset into study-level report cards

We developed a pipeline to automatically generate individualized report cards based on the trial transparency dataset (see Supplement 4 Figure 1). In the first step, we designed a template of the report card in Affinity Designer, a vector graphics editor. The report card template is a visual representation of a trial’s performance across transparency practices. Since trials perform differently across transparency practices, the report card template includes all possible outcomes of a trial’s performance represented as layers. The report card template was exported as Scalable Vector Graphics (SVG), which is an image format based on Extensible Markup Language (XML). This allows the report card template to be accessed and modified using code. In the second step, report cards for each trial were assembled automatically using a custom-made Python script that selects the correct layers to include for each trial based on the trial transparency dataset. In the third step, each report card was automatically exported as a PDF for dissemination to study participants. An example report card is displayed in Figure 1 (a). The infosheet (Figure 1 (b)) was generated in Affinity Designer and Affinity Publisher and adapted on an iterative basis.

### Feasibility study

We conducted a feasibility study to pilot the use of the tools at the Charité – Universitätsmedizin Berlin, in collaboration with the institutional Clinical Study Center. The study used a cross-sectional, interventional design and included a descriptive analysis of two main outcomes: self-reported evaluation of the tools via a survey, and improvement in transparency practices following the intervention of the tools. All trials received the intervention (single-arm), as the number of trials eligible to be included in our analysis was too small for a control group and sufficient power to detect a minimum effect size of interest.

#### Ethical approval and study protocol

This study was approved by the ethics review board of the Charité – Universitätsmedizin Berlin (#EA1/337/21). The study protocol was preregistered and is openly available in the Open Science Framework (OSF) at: https://doi.org/10.17605/osf.io/stnp5. Deviations to the protocol are outlined in OSF: https://osf.io/wxayg.

#### Sample development

This study used a cohort of trials and associated investigators. Screening criteria were assessed for both trials and investigators, in part prior to study launch and in part prior to analysis. Criteria are outlined below and detailed in Supplement 2 Table 2.

##### Trials

This study draws from two cohorts of registered clinical trials and associated results, referred to as “IntoValue” (Riedel et al., 2021). The IntoValue dataset consists of clinical trials led by a German UMC (i.e., as either sponsor, responsible party, or host of the principal investigator), registered in ClinicalTrials.gov or the German Clinical Trials Register (DRKS), and considered as complete per the registry between 2009 and 2017. For this feasibility study, the IntoValue dataset was limited to trials led by the Charité with a completion date between 2014 and 2017 (i.e., the second IntoValue cohort); these more recently completed trials were selected, given the higher likelihood of reaching investigators and increased feasibility of implementing practices. To reflect the most up-to-date status of trials, we downloaded updated registry data for the trials in this cohort on 19 May 2022 and reapplied the original IntoValue exclusion criteria: study completion date before 2014 or after 2017, not considered as complete based on study status, and not interventional. Trials were excluded if no Charité investigators were found or reached.

##### Trial investigators

We developed a list of investigators for our sample of trials based on contact information retrieved from clinical trial registries. We used the following registry fields: from ClinicalTrials.gov, responsible party (if this included principal investigator), and overall study officials (principal investigator, study director, study chair); from DRKS, primary sponsor, and contacts for scientific and public queries. As contact information is often removed from registrations after trial completion (O’Neill et al., 2014; Viergever et al., 2014), we used the R package 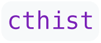 (Carlisle, 2022) to retrieve the current (as of 19 May 2022) as well as all historical versions of registrations. If no contact information was available in the registry, we searched Google and the Charité address book using the investigator’s name in the registry and the Charité email pattern, as well as trial details such as trial registration number and title keywords as needed. If multiple investigators or multiple emails for the same investigator were available, all were used; if the registration provided an email for the project or research group, this was included in addition to the investigator email(s). Investigators were excluded if no contact information was found, and trials were excluded if no contact information for any trial investigator was found. Per our preregistration, after the email-based intervention, investigators were excluded if all emails were undeliverable or if auto-replies indicated extended leave throughout our study timeframe; trials were then excluded if no trial investigator remained.

#### Materials

##### Transparency practices and report cards

In line with the requirements for the report cards (see “Trial transparency dataset”), the IntoValue trial cohort includes data on transparency practices. Data obtained through automated methods were updated at several time points during the study. Registry data for ClinicalTrials.gov was queried from the AACT (Clinical Trials Transformation Initiative, n.d.), and DRKS was webscraped prior to the start of the intervention and at the end of follow-up. At study end, we additionally used 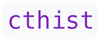 (Carlisle, 2022) to get historical versions of registrations in order to complete the analysis of each study time point. To determine the open access status, we queried the Unpaywall API (using UnpaywallR (Riedel and Franzen, 2022)) at each study time point; to determine publisher permissions for self-archiving, we queried the Shareyourpaper API (using a custom-made Python script (Franzen, 2023)) at study launch. Additional manual validations were done for the open access status of publications as well as for cross-registration in the EUCTR. Protocols for these validations were prospectively uploaded on OSF (https://osf.io/3upgy and https://osf.io/mjpzh), and methods are summarized in Supplement 5. The resulting trial transparency dataset was used to generate the report cards. After study launch, we found a bug in the code and subsequently made 6 amendments to the dataset and report cards, namely we added: 1 cross-registration in EUCTR, 4 publication links in DRKS, and 1 summary result in DRKS. More detailed information can be found in the project repository in GitHub: https://github.com/quest-bih/trackvalue.

##### Questionnaire development and pretesting

We conducted an anonymous survey to assess the usefulness of the report cards and infosheet. The survey was generated in LimeSurvey and hosted on a server at the QUEST Center for Responsible Research at the Berlin Institute of Health at Charité – Universitätsmedizin Berlin. After informed consent and prior to starting the survey, respondents were asked whether they had reviewed the materials, and if not, they were prompted to do so and return to the survey. Respondents were asked to evaluate the report card and infosheet using a 5-point Likert scale across four items for each tool. Respondents were also asked to indicate their role(s) in the trial, and were invited to provide corrections to the report card(s), if applicable, as well as general comments. The survey was presented in German. The design of the survey was shaped by feedback obtained in cognitive interviews (see Supplement 1). The survey is provided in OSF (https://osf.io/5z463).

##### Administration of the intervention and survey

Investigators were sent an invitation email including: a letter (co-signed by the Charité Clinical Study Center) listing all their trials, a report card for each trial they led, and an infosheet. The email and letter also included an invitation to participate in the anonymous survey as well as a notice that the transparency of their trials would be re-evaluated 3 months after the first email (see a sample invitation in OSF: https://osf.io/9qkdp). Emails were sent from a functional mailbox created for the study that was regularly monitored over the course of the study. Any auto-replies indicating undeliverable emails or extended leave were preserved. Follow-up emails were sent at weeks 1, 5, and 11. As the survey was anonymous, and we did not know who had already completed it, all reminders were sent to all investigators included in the study. We reviewed the survey responses on an ongoing basis to identify any valid corrections to the report cards provided by investigators. For valid corrections, we updated our dataset and generated and sent amended report cards to all investigators associated with the trial.

#### Analysis

##### Analysis of perceptions of the report card and infosheet

We describe investigators’ perception of the report card and infosheet per the survey. Our primary outcome is how much respondents agree with four positive statements about each tool. Statements (for example, “The infosheet is clear.”) were presented with a 5-point Likert-type response scale, with options ranging from 1 = strongly disagree, to 5 = strongly agree. We report the proportion and means with 95% confidence intervals of each Likert response. Survey responses were considered complete if all Likert items were completed, and as all Likert items were required, we did not evaluate missingness. We report the survey response rate, the response timeline across the survey fielding period, and exclusion from analysis at each stage (e.g., due to undeliverable emails). As the survey was anonymous, we could not evaluate non-response error or adjust for possible non-representativeness of the respondents; we could not determine whether unique respondents came from unique visitors. We evaluated response time as a quality check for active participation and to avoid counting bots. We report the number of respondents who reported not reviewing materials before starting the survey and perform a subgroup analysis of these respondents. We also perform a subgroup analysis of study leadership. We report the number of corrections, and evaluate and report the number which are valid; corrections were reflected in subsequent report card versions. We also report the number of comments, after removing non-responses and describe common themes identified in respondent feedback. Corrections and comments were reviewed and adjudicated by consensus by two researchers (MSH a

##### Analysis of improvement in transparency practices

We assessed trials for three transparency practices that could still be changed after trial completion: 1) reporting of summary results in the registry, 2) publication links in the registration, and 3) open access status of publication. The publication-based analyses were limited to trials with publications available at the time of intervention launch; the open access analysis was further limited to trial publications that could be self-archived in an institutional repository based on publisher policies (“green open access”) at the time of intervention launch. We report the overall proportion of trials performing each practice at the following time points relative to our intervention: 3 months prior to launch, at launch, 3 months after launch. As a protocol deviation, we additionally report rates a year after the intervention.

### Software, code, and data

All the code generated in this study is openly available under an open license in GitHub: https://github.com/quest-bih/trackvalue. Data collection was conducted in Python [Version 3.9] and R [Version 4.3.2], and all analysis was conducted in R. This manuscript was written by interleaving regular prose and analysis code in Quarto [Version 1.4.549]. With the exception of investigator contact information and survey comments and corrections, all the data is openly available in the GitHub repository of the project and Zenodo: https://doi.org/10.5281/zenodo.10467054.

### Reporting guidelines

This study uses both an intervention and a survey, and follows the CROSS Consensus-Based Checklist for Reporting of Survey Studies (Sharma et al., 2021) and CONSORT for Pilot and Feasibility Trials (Eldridge et al., 2016). Reporting checklists are available in Supplement 6.

## Discussion

We developed scalable, open source tools to provide individualized feedback and guidance on research transparency across phases of a clinical trial. We investigated the feasibility of using these tools to improve clinical trial transparency in a sample of completed trials led by researchers at the Charité – Universitätsmedizin Berlin, one of the largest university hospitals in Europe (https://www.charite.de/en). Overall, researchers found the tools helpful to inform them of the transparency of their trial and to raise awareness of relevant guidelines, laws, and resources. However, improvement across transparency practices one year after dissemination of the tools was limited. This study demonstrates the feasibility of generating and communicating study-level feedback to researchers, and can inform future interventions to improve transparency and the adoption of responsible research practices more broadly.

### Summary of findings

#### Perceptions of the report card and infosheet

Survey responses indicated that the report cards and infosheet were generally well received by study participants. The infosheet, in particular, stood out as being a valuable resource, suggesting that succinct visual summaries of what is expected of investigators (based on applicable laws, guidelines, and policies) may be a fruitful approach to build awareness of transparency requirements alongside other forms of training. Participants’ views on the report cards were largely positive. Lower ratings on the usefulness of the report cards to improve trial transparency may reflect the need for additional support or the removal of barriers to implement changes in practice. We attempted to bridge this gap by linking to detailed guidance on how to perform each practice.

#### Transparency improvements in practice

Trial transparency improved minimally over the follow-up period, with improvements largely limited to linked publication in the registry. The discrete improvement for this practice, which involves manually adding a link to a results publication in the registration, may reflect the lower time and effort needed compared to other practices, or perceived hurdles associated with summary results reporting (e.g., fear of jeopardizing journal publication (Bruckner, 2019)) and self-archiving (e.g., fear of copyright infringement (Swan and Brown, 2005)). Furthermore, our study called for action on trials reported as complete in the registry up to 8 years prior to this intervention, which may have been too long ago for these practices to be feasibly addressed. Prospective reminders while a study is ongoing or for recently completed studies, as already implemented in several registries (e.g., ISRCTN (Taylor, 2019)), may be more effective in driving change.

Perceived barriers to implementing transparency practices were noted in investigators’ survey responses. While the survey did not aim to solicit feedback beyond the tools, numerous respondents volunteered several challenges (see Supplement 3 Table 1), including lack of clarity around best practices (e.g., applicable guidelines and/or laws in case of multiple registrations of a single trial), diffusion of responsibility (e.g., limited control over transparency practices due to journal guidelines), and obstructive institutional processes and bureaucracy around the conduct of trials, which take time and resources away from other activities. Taken together, investigators’ positive perception of the materials yet limited improvement in transparency practices, as well as unprompted comments on challenges, suggests that implementing research transparency is a multifaceted issue and that interventions likely need to be embedded within systemic changes.

### Research in context

Previous empirical research has explored the impact of interventions at the level of individual studies to increase clinical trial transparency. Maruani and colleagues conducted a randomized controlled trial in 2014 to evaluate the impact of sending email reminders to investigators of completed Phase IV trials about their requirements to post results (Maruani et al., 2014). At six months following their intervention, results had been posted for 24% of trials that had received the intervention versus 14% in the control group, suggesting that reminders may effectively prompt results reporting. A 2023 study at the German registry DRKS similarly found that email reminders were effective in prompting investigators to complete draft registrations: 5 weeks after the intervention, 11% of trials that had been sent an email had completed their registrations, compared with 4% of trials that received no communication (Bieselt, 2023).

Approaches such as report cards or reminders may have greater impact when embedded within a holistic behavior change approach that brings together capability, motivation, and opportunity (Michie et al., 2011), as suggested by previous work across research, policy, and practice. For example, the launch of the EU Trials Tracker as a public audit tool was complemented by an advocacy campaign, which eventually spurred policymakers and other stakeholders to demand change from institutions (Lamb, 2019). Research performing institutions subsequently rapidly improved their capacities to address these practices, and within a few years, universities policies had been strengthened (Keestra et al., 2022), and trial reporting increased from 50% in 2018 to 84% in September 2023 (Bennett Institute for Applied Data Science, n.d.). Additional external factors further reinforced opportunity: in February 2020, the Make It Public strategy was adopted in the United Kingdom, whereby clinical trials are automatically and centrally registered by the Health Research Authority following ethics approval, which saves researchers this extra step (Health Research Authority, 2020).

In the United States, efforts to understand existing support structures for trial registration and reporting (Mayo-Wilson et al., 2018) were accompanied by improvements in transparency at academic organizations. Institutional programs to support compliance with registration and reporting requirements launched a variety of activities, including audits to identify problem records and bring them into compliance, prospective reminders, policy development, enforcement, standard operating procedures, centralized resources, training, and consultation (Keyes et al., 2021; O’Reilly et al., 2015; Snider et al., 2020). Following the implementation of such a program at Johns Hopkins University School of Medicine, non-compliance for mandated registration and reporting dropped from 44% (339/774) in 2015 to 2% (32/1,304) in 2020 (Keyes et al., 2021). More broadly, the emergence of interventions implemented by key stakeholders at different levels offers a rich opportunity to investigate effective, multifaceted strategies for research improvement.

### Strengths, limitations, and challenges

Our study has several strengths. The report cards offer a single resource that brings together individualized feedback and guidance on the implementation of specific practices. The approach to generate the report cards is openly available and can be scaled to other institutions and contexts that have, or are seeking to develop, an overview of their studies and associated practices. Importantly, this approach leverages automated approaches to drive transparency at scale, while not being tied to a specific stakeholder or infrastructure. Moreover, we collaborated with an institutional core facility: this approach may be a strength by increasing institutional buy-in and the likelihood of sustainable adoption of such a tool; however, it may also have shifted investigators’ attention away from the tools towards institutional aspects.

Our study also faces several limitations. The report cards rely on the information in the registry being accurate and up-to-date, which is not always the case (DeVito and Goldacre, 2022; Fleminger and Goldacre, 2018; Viergever et al., 2014). For EUCTR trials, prospective registration was assessed based on dates available in the registry; however, in principle all EUCTR trials should be prospectively registered, as registration is linked to regulatory approval by national competent authorities. The small sample size in this study precluded randomization and a control group with sufficient power to detect a minimum effect size of interest. The design therefore did not allow us to test whether our intervention drove the changes (i.e., causality), which may also have been prompted by researchers themselves or other co-occurring pressures in the German clinical trial landscape (Bruckner, 2022). However, as our aim was to test the feasibility of delivering such an intervention, the lessons learned may inform a larger randomized controlled trial. Another limitation of our study is that the invitation letter and survey were only provided in German, while the report card(s) and infosheet were provided in English. This may have limited the reach or understandability of the materials. Future studies may consider generating multilingual materials.

We identified several challenges related to material development and the intervention, which may impact the scalability of this type of intervention. Despite the use of automated approaches, additional manual checks were still required to validate the data. Another challenge was balancing comprehensiveness and clarity in the infosheet; this could be supported by engaging visual design experts and by using dynamic formats such as HTML instead of static PDFs. Additionally, step-by-step instructions on how to implement transparency practices were not available across all registries at the time of the intervention. Contact information of investigators was not always readily available in the registry, and in some cases required searching historical versions of a trial or internet searches.

### Scaling beyond transparency in clinical trials

The report cards were designed for scalability and extensibility. They may be adapted to provide individualized feedback and guidance on further transparency or responsible research practices, such as data sharing, reporting of robustness measures (e.g., randomization), measures to improve patient-orientedness (e.g. patient engagement in study design), or reporting of ethical safeguards (e.g., conflict of interest disclosure). The report cards may also be adapted for use in other types of studies and disciplines. This requires a core set of practices to monitor, an overview of studies, and automated and/or manual approaches to extract, process, and validate the data to include in the report cards (Weissgerber et al., 2021). While many study types and disciplines do not yet have formalized best practices for responsible research comparable with those for clinical trials, a core set of practices to monitor within a discipline may be established through community consensus, as done in biomedicine (Cobey et al., 2023). We expect broader applicability of tools that provide study-level feedback, as registration becomes more common across certain fields (e.g., animal research (Heinl et al., 2022)) and serves as a source of study information complementary to publications.

### Implications and future work

Future work is needed to further evaluate the use of report cards to address transparency, and iteratively transfer this potential solution to policy and practice. Our feasibility study may inform a larger randomized controlled trial at the level of multiple institutions, and institution-led behavior change interventions more broadly. These studies may consider following up with investigators who implement practices to understand whether the intervention indeed prompted the behavior. Future work may also consider evaluating prospective use of the tools: report cards that prompt investigators before a specific practice is due; an infosheet disseminated at study start (e.g., ethics approval) that sets out expectations for researchers before the work is conducted. The report cards may be particularly well suited to help funders monitor and support compliance with grant requirements, which typically span the entire project lifecycle.

Future studies should adopt a systemic approach and consider the role of capability, motivation, and opportunity when designing an intervention to improve transparency (Michie et al., 2011). For example, combining the report cards with initiatives that reinforce motivation, such as awards or broader recognition of responsible research practices in research assessment (Moher et al., 2020), may be an effective approach. Proposed solutions will also need to be tailored to the specific context and resources of a given institution. Engaging with researchers and other stakeholders, who have first-hand experience of challenges and needs, will be key towards identifying actionable areas for improvement to drive sustainable change.

## Data Availability

https://github.com/quest-bih/trackvalue

## Acknowledgements

We would like to thank the Clinical Study Center of the Charité – Universitätsmedizin Berlin for supporting this intervention. We are grateful to the participants of our cognitive interviews as well as Sarah White and Tony Keyes from the US-based Clinical Trials Registration and Results Reporting Taskforce for their valuable feedback on the report cards and the infosheet. Many thanks to Benjamin Gregory Carlisle for assisting us with the automated email invitations and reminders, Tamarinde Haven and Christiane Wetzel for their input on the survey design, Susan Abunijela for support with performing manual checks of the open access status, Samruddhi Yerunkar for performing manual checks of cross-registrations and visualizing the survey responses, Nicole Hildebrand for manually searching for investigator contact information not available in the registries, as well as Martin Holst, Martin Haslberger, and Friederike Kohrs for their support with language translation. We also thank the Service Unit Biometrics of the Institute for Biometrics and Clinical Epidemiology (iBikE) of the Charité – Universitätsmedizin Berlin for consulting on the statistical analysis. Finally, we would like to thank those who gave their time to participate in our study.

## Declarations

The authors declare no competing interests.

## Author Contributions

Delwen L Franzen: Conceptualization, Data Curation, Formal Analysis, Investigation, Methodology, Project Administration, Resources, Software, Validation, Visualization, Writing - Original Draft, Writing - Review & Editing; Maia Salholz-Hillel: Conceptualization, Data Curation, Formal Analysis, Investigation, Methodology, Project Administration, Resources, Software, Validation, Visualization, Writing - Original Draft, Writing - Review & Editing; Stephanie Müller-Ohlraun: Conceptualization, Methodology, Writing - Review & Editing; Daniel Strech: Conceptualization, Funding Acquisition, Methodology, Supervision, Writing - Review & Editing.

## 1 Expert reviews and cognitive interviews on the materials and survey

### Methods

We solicited expert reviews to get feedback on the content and design of the invitation letter, report cards, infosheet, and survey. We contacted content experts, experts in questionnaire development, and experts in statistical analysis (Dillman et al., 2014, Guideline 7.7). Reviews were unstructured and tailored to each person’s expertise. Content experts included researchers and administrators involved in the conduct and management of clinical trials. Colleagues with extensive experience conducting surveys reviewed and consulted on the questionnaire. A statistical consultant at the Charité reviewed the analysis plan prior to preregistration of the protocol.

We also performed think-aloud cognitive interviews to identify challenges with the visual design, wording, and navigation of the materials, and to evaluate whether the respondents interact with the materials and understand the questions as intended by the research team (Dillman et al., 2014, Guideline 7.8). Participants (n = 3) were identified within our networks and were selected based on previous or ongoing experience conducting or administering clinical trials at the Charité, in order to maximize similarity with the sample population. Participants received an invitation email to participate in a 1-hour interview conducted over Microsoft Teams. One team member led the interview (MSH) while another team member (DLF) observed and made notes of the interview. After informed consent, participants were asked to walk through the invitation letter, report card, infosheet, and survey and think aloud to capture their thought processes when engaging with the study materials. The cognitive interview guide is available in OSF (https://osf.io/xtpjc). Based on the feedback shared in each interview, we modified the materials prior to the following interview.

### Results

Themes (Table 1) for the design of the report card included: provide actionable information by combining feedback and guidance, and by highlighting practices that can still be improved; contextualize feedback and guidance by providing study details and the normative basis for each practice; use clear visuals and accessible language to convey the message quickly. The main theme for the design of the infosheet was balancing comprehensiveness and clarity, by tailoring the information to the audience (e.g., acknowledge institutional guidelines and resources), and by carefully considering the structure and layout. With a few exceptions due to feasibility, the feedback was incorporated into the report cards and infosheet.

**Table 1:** Emerging themes for the design of the report cards and infosheet developed in this study.

| Theme | Notes |
| --- | --- |
| Provide actionable information | • Combine feedback and guidance for improvement in the same place<br>• Limit guidance to practices that can still be improved<br>• Include resources to reduce barriers to taking action (e.g., hyperlink to implement change in registry)<br>• Link to institutional support, where applicable (e.g., core facilities)<br>• Use language to emphasize actionability (e.g., “You can still improve your trial’s transparency”) |
| Visual representation of performance | • Convey the right message quickly using appropriate symbols and colors<br>• Use colorblind-friendly colors |
| Contextualize feedback and guidance | • Highlight the normative basis for each practice (e.g., ethical guideline or legal requirement)<br>• Provide study details for each practice to contextualize feedback (e.g., days between trial completion and results publication)<br>• Provide study details in a recognizable way (e.g., consider whether to refer to a publication with a citation or a publication title) |
| Accessible Language | • Avoid using acronyms. If acronyms are used, check for familiarity with the acronym (e.g., EudraCT or EUCTR to refer to the EU Clinical Trials Register)<br>• Avoid language that may be perceived as established terminology (e.g., “trial score”)<br>• Use simple and concise language<br>• Use specific language (e.g., “earliest publication” if report card only includes information about the earliest publication)<br>• Check for possible misunderstandings |

Report card
| Theme | Notes |
| --- | --- |
| Balance comprehensiveness and clarity | Specify that the infosheet includes generic recommendations and is not study-specific<br>Tailor the information in the infosheet to the audience: where applicable, align recommendations with institutional or funder policies, and link to existing support infrastructure (e.g., institutional core facilities)<br>Where applicable, provide general information beyond specific practices (e.g., in which registry(-ies) should a trial be registered?) |
| Structure and layout | Carefully consider how to structure the infosheet (e.g., aggregate resources across all practices in a single place, avoid placing the abbreviations table at the bottom, so that it is not missed)<br>Arrange practices in a meaningful way (e.g., according to where they are to be implemented, or guidelines vs. regulations)<br>Use hyperlinks to other sources to provide additional information<br>Limit the infosheet to 1 page<br>Consider using a landscape layout for better readability |
| Language | Provide an abbreviations table if using acronyms |

## 2 Screening criteria and characteristics of trials and investigators

**Table 1:** Characteristics of included trials and investigators.

|  |  |
| --- | --- |
| **Investigator characteristics** | **N = 92** |
| Contact email found in registry<sup>1</sup> | 54 (59%) |
| **Trial characteristics** | **N = 155** |
| Registry |  |
| ClinicalTrials.gov | 140 (90%) |
| DRKS | 15 (9.7%) |
| EUCTR<sup>2</sup> | 28 (18%) |
| Completion status |  |
| Completed | 129 (83%) |
| Terminated | 12 (7.7%) |
| Unknown | 14 (9.0%) |
| Completion year |  |
| 2014 | 38 (25%) |
| 2015 | 50 (32%) |
| 2016 | 40 (26%) |
| 2017 | 27 (17%) |
| **Trial transparency practices included in report cards** |  |
| Prospective registration | 94 (61%)<sup>3</sup> |
| ClinicalTrials.gov | 90 (64%) |
| DRKS | 4 (27%) |
| EUCTR<sup>4</sup> | 27 (96%) |
| Summary results in registry | 16 (10%)<sup>3</sup> |
| ClinicalTrials.gov | 16 (11%) |
| DRKS | 0 (0%) |
| EUCTR | 20 (71%) |
| Summary results in registry within 1 year of completion | 8 (5.2%) |
| ClinicalTrials.gov | 8 (5.7%) |
| DRKS | 0 (0%) |
| Results published in journal article | 102 (66%) |
| Results published in journal article within 2 years of completion | 54 (53%) |
| Trial registration number in publication abstract | 34 (33%) |
| Trial registration number in publication main body | 58 (57%) |
| Publication open access<sup>5</sup> | 68 (67%) |
| Publication linked in registration | 53 (52%)<sup>3</sup> |
| ClinicalTrials.gov | 47 (53%) |
| DRKS | 6 (43%) |
| **Trials eligible for each transparency practice** |  |
| Summary results in registry | **N = 155** |
| Completed prior to launch | 16 (10%) |
| Not completed prior to launch | 139 (90%) |
| Publication linked in registration<sup>6</sup> | **N = 102** |
| Completed prior to launch | 53 (52%) |
| Not completed prior to launch | 49 (48%) |
| Publication open access<sup>7</sup> | **N = 100** |
| Completed prior to launch | 68 (68%) |
| Not completed prior to launch | 32 (32%) |

<sup>1</sup> We checked both the current (as of 19 May 2022) and historical versions of registrations.
<sup>2</sup> EUCTR was not used as a primary registry data source. Hence, included EUCTR trials reflect cross-registered trials. within the 155 trials with a registration in ClinicalTrials.gov or DRKS.
<sup>3</sup> Limited to trials registered in ClinicalTrials.gov or DRKS, the primary registry data sources in this study. EUCTR is not included.
<sup>4</sup> Prospective registration was assessed based on dates available in the registry. However, in principle all EUCTR trials should be prospectively registered, as registration is linked to regulatory approval by national competent authorities.
<sup>5</sup> Open access includes gold, hybrid, bronze, and green.
<sup>6</sup> Limited to trials with a publication.
<sup>7</sup> Limited to trials with a publication and a publisher permission to self-archive the paper in an institutional repository. For two closed access publications, we could not determine whether such a permission was available.

**Table 2:** Trial and investigator inclusion criteria.

| **Timepoint** | **Criteria** |
| --- | --- |
| Prior | Included the Charité as the sponsor, responsible party, or host of the principal investigator |
| Prior | Study completion date between 2014 or after 2017, study status considered as complete, and interventional |
| Prior | One or more investigator with email |
| Post | One or more investigator with successfully delivered email and no auto-reply indicating extended leave throughout our study timeframe |
| Trial |  |
| **Timepoint** | **Criteria** |
| Prior | In ClinicalTrials.gov responsible party (if this included principal investigator), overall study officials (principal investigator, study director, study chair), and central contact person and backup, or in DRKS primary sponsor and contacts for scientific and public queries |
| Prior | One or more emails found in registry or Google search |
| Post | Not all emails were undeliverable and no auto-replies indicated extended leave throughout our study timeframe |
| Investigator |  |

## 3 Survey administration and detailed results

### Response timeline across the survey fielding period

**Figure 1:**
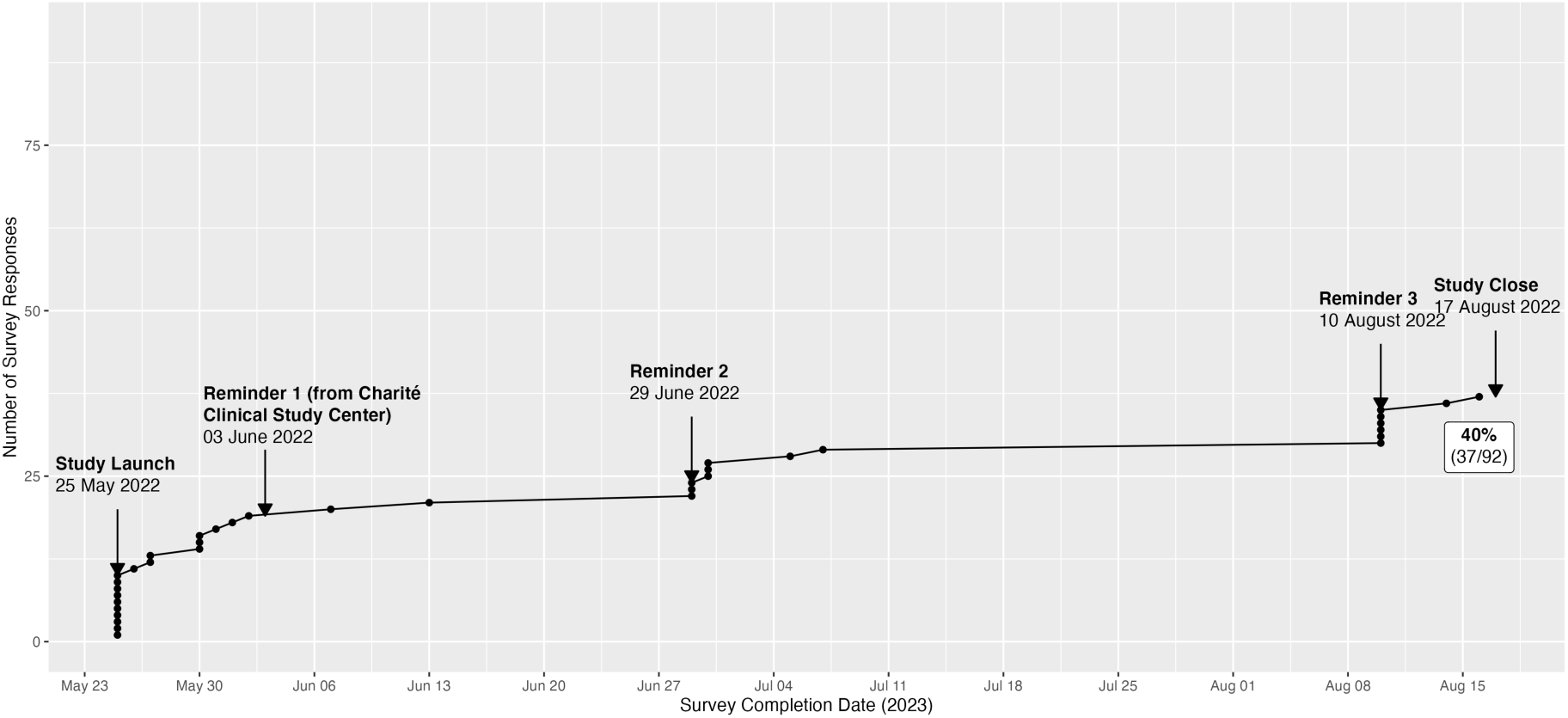
Timelines of survey responses and key survey dates. The first reminder was sent on behalf of the Clinical Study Center and prompted investigators to review the initial invitation email but did not contain a link to the survey or any other materials. Reminders 2 and 3 both included a link to the survey as well as the report card(s) and infosheet as attachments.

### Survey respondents roles

**Figure 2:**
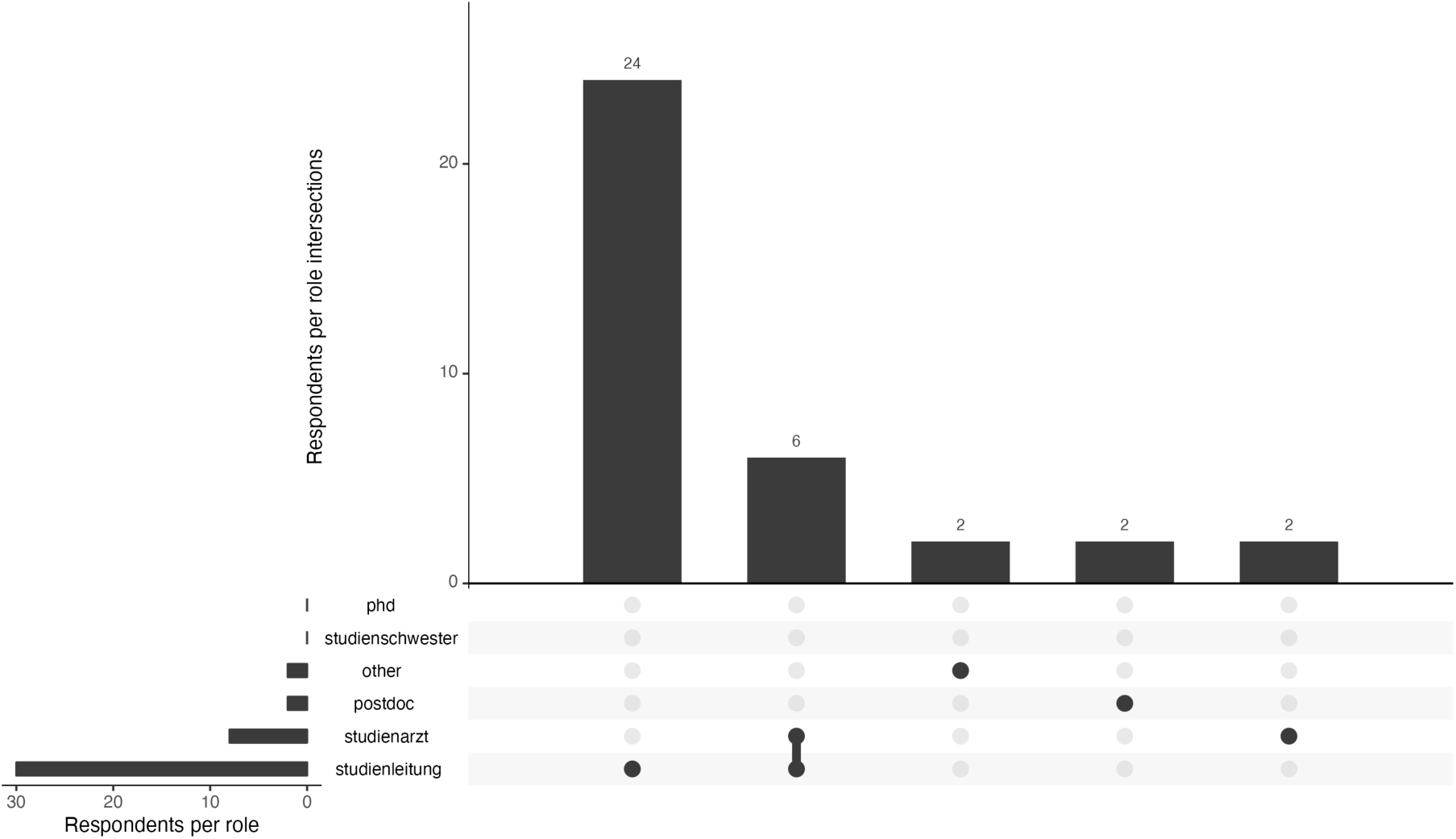
Survey respondents self-reported roles in trials.

The majority of respondents self-reported as study leads (n = 24, 65%), while an additional 6 (16%) self-reported as both study leads and doctors. The remaining respondents identified as either doctors, postdocs, or another position (per group, n = 2, 5.4%).

### Themes of comments provided by respondents

**Table 1:** Summary of comments from the survey and emails.

| Suggestions for improvement on the materials | Detail |
| --- | --- |
| Scope | • Consider including all results publications associated with a trial<br>• Consider expanding report cards to other types of trials<br>• Interest in further information on required documentation for different types of studies<br>• Simplify and focus on essential aspects |
| Visual design | More professional graphics and design of the report cards and infosheet |
| Implementation | Send report cards at regular intervals across a study lifecycle |
| Accessibility | Make the survey bilingual |
| Times for results reporting | Use event-based times instead of fixed times for results reporting |

| Challenges related to implementing transparency | Detail |
| --- | --- |
| Institutional barriers | • Management and conduct of clinical trials characterized by obstructive, time-consuming institutional processes<br>• Report cards seen as additional administrative bureaucracy, which takes time and resources away from other things<br>• Insufficient protected time for publishing |
| Legitimacy | Unclear authority of the research team to evaluate studies |
| Diffusion of responsibility | • Limited control over transparency practices due to journal guidelines<br>• Concern about non-reporting of summary results before publication in a journal |
| Lack of clarity on practices | • Perception that in case of multiple registrations of a trial, practices can be implemented in only one registry<br>• Focus on legal requirements<br>• Publication abstract or linked publication instead of summary results |
| Technical difficulties | Technical challenges accessing/managing registry entries |

### Trial investigator corrections to the report card

Investigators had the opportunity to provide corrections to their report card. Respondents suggested 18 corrections on 11 trials across the survey and emails. In our manual review, we found that many were in fact not corrections but rather incorrect understandings of the practices or additional information on the trials beyond the scope of the report card. This, for example, included links to additional results publications, or information that a publication was open access when in fact it was only accessible via an institutional subscription. After limiting these to valid corrections, 5 corrections regarding 4 trials remained: 1 missed earlier results publication, 1 missed results publication, 1 missed cross-registration in EUCTR, and 2 missed publication links in DRKS.

### Subgroup analyses of survey responses

We conducted two post-hoc subgroup analyses. Figure 3 shows means and 95% confidence intervals for agreement (i.e., Likert-type response item) with each of eight statements about the report card and materials, across all respondents as well as the for the subgroup analyses.

#### Study role

We explored perceptions of the report card and infosheet across different study roles. We did not observe a meaningful difference between study leads and doctors (n = 32) versus other respondents (n = 5), who were a smaller group with a wide spread of responses.

#### Review of report card and infosheet

A total of 3 respondents reported not reviewing the report card and infosheet prior to starting the survey and were asked to do so prior to continuing the survey. Per our prespecified inclusion criteria, we did not exclude these responses. However, response time did not significantly differ between those who were and were not asked to review the materials (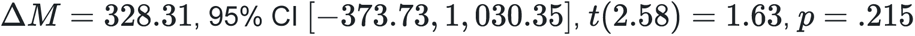), which suggests that they did not pause the survey to review the materials. We therefore conducted a post-hoc subgroup analysis. Perceptions of the report card and infosheet were substantially lower among those who reported not reviewing the materials prior to starting the survey: all Likert responses across all 8 items for all 3 respondents were the most negative option, and their free-text responses also included negative comments.

**Figure 3:**
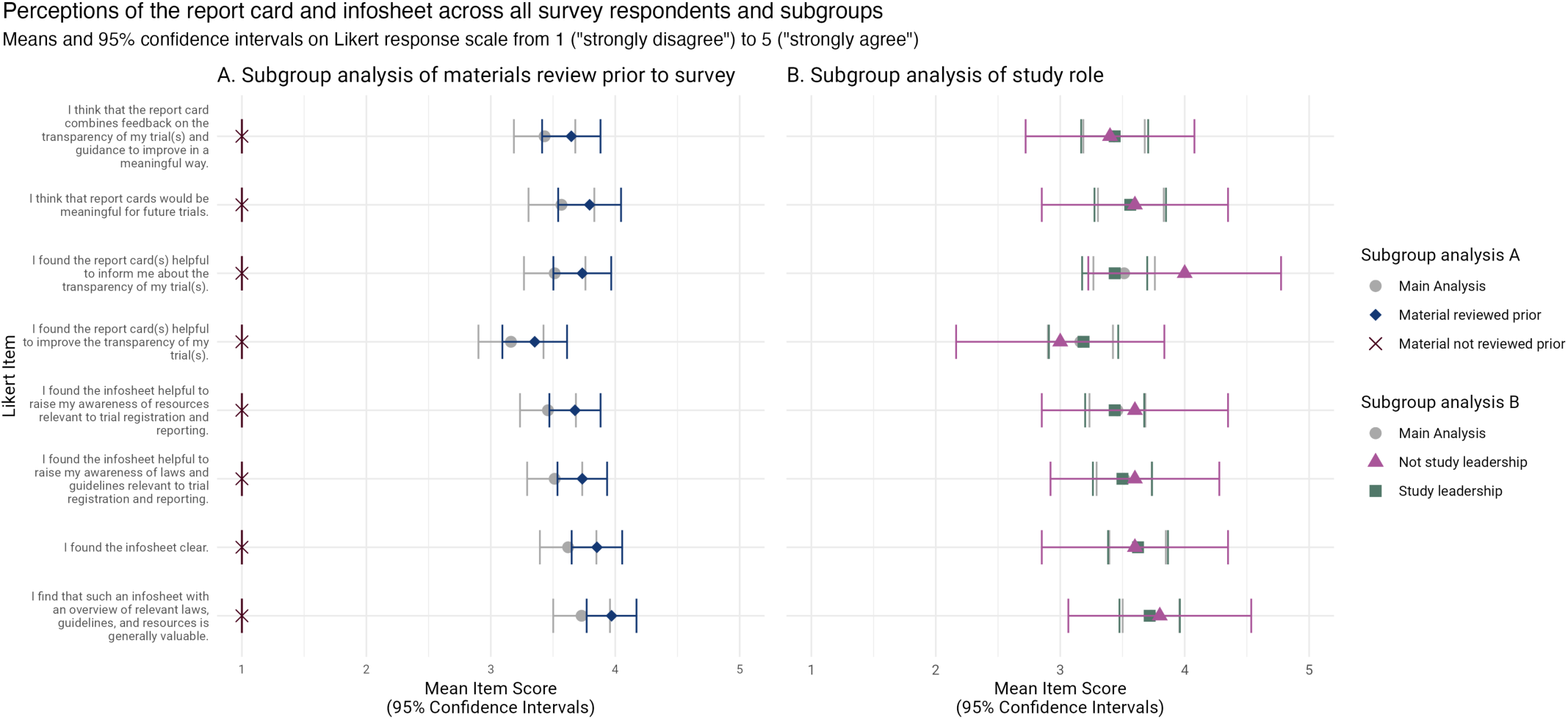
Perceptions of the report card and infosheet across all survey respondents and subgroups.

## 4 Steps to generate individualized trial transparency report cards

### Main steps to prepare a trial transparency dataset for use with the report cards

**Table 1:** Steps to create a trial transparency dataset for use with the report card tool.

| Step | Manual or automated | Notes |
| --- | --- | --- |
| Generate cohort of trials at the level of an institution | Semi-automated | Trials associated with a given institution may be extracted from the registry using automated methods. However, a manual review of affiliations is still required. |
| Find results publication(s) associated with trials | Semi-automated | Linked results publications may be extracted from the registry using automated methods. However, a manual search for results publications is required since: a) linked publications in the registry are not always results publications (e.g., protocols, reviews), and b) results publications are often not linked in the registration. |
| Assess transparency of trials (e.g., prospective registration) | Automated to semi-automated | Transparency practices can be evaluated using code, depending on the registry. |
| Data validation (e.g., cross-registrations) | Manual | Some elements of the report card may require additional manual validation, such as for example suspected cross-registrations. |

Additionally, the following code repositories provide more detailed information (e.g., codebook) on the automated steps used to generate components for the report cards:

- https://github.com/quest-bih/trackvalue
- https://github.com/maia-sh/intovalue-data

### Automated pipeline to generate individualized report cards

The pipeline is based on a report card template that includes all possible outcomes of a trial’s performance represented as layers. The report card template is exported as SVG, which is an image format based on XML. This means that the report card template can be accessed and modified using code. A custom-made Python script selects the correct layers to include for each trial based on a trial transparency dataset. Each report card is automatically exported as PDF.

**Figure 1:**
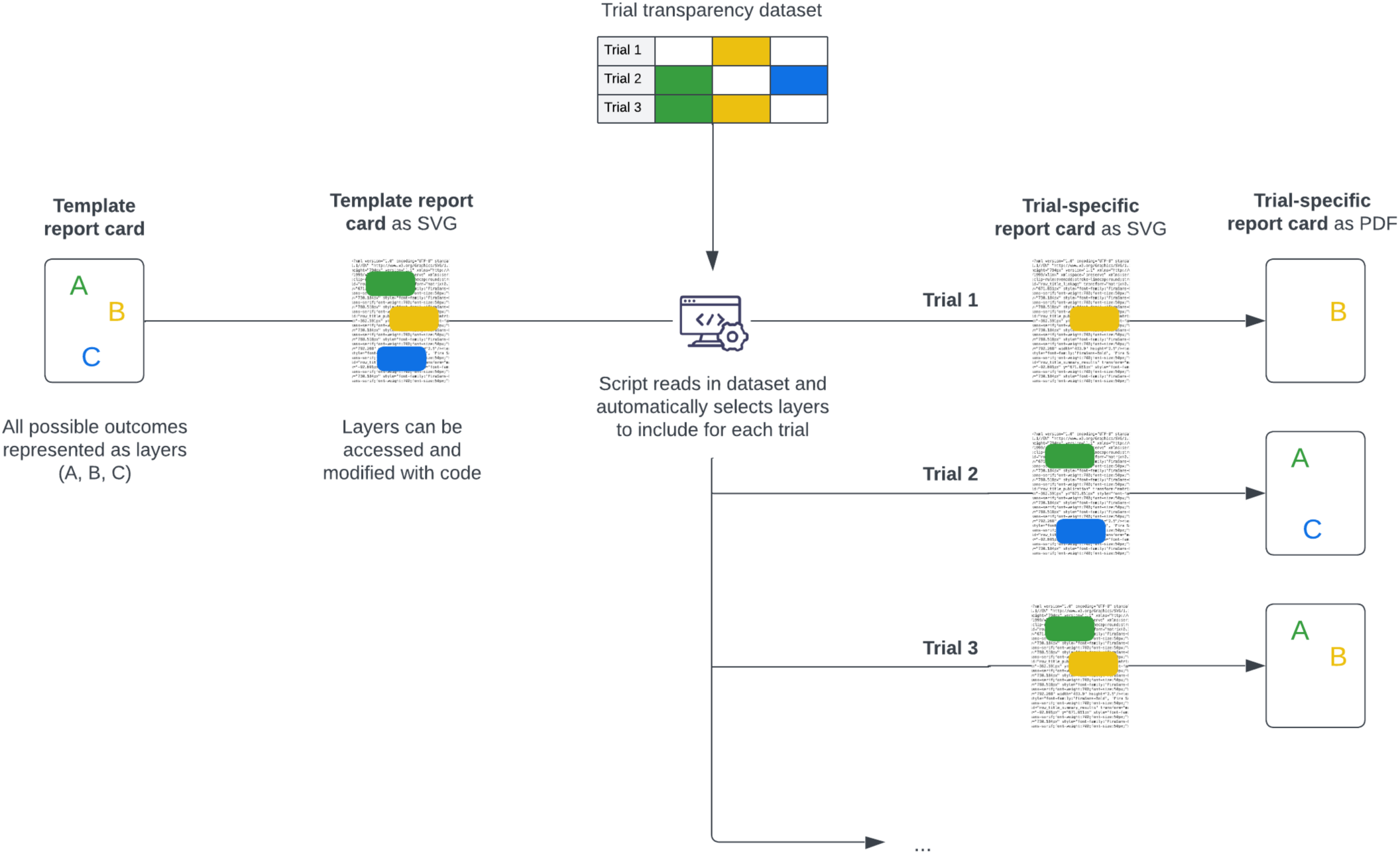
Automated pipeline to generate individualized report cards based on a trial transparency dataset. PDF: Portable Document Format; SVG: Scalable Vector Graphics.

## 5 Methods for manual validation of open access status of publications and cross-registration in the EUCTR

The report cards developed for this study included references to any known EUCTR cross-registration (additional registrations of a single trial in a different or same registry). To best ensure that our dataset accurately captured cross-registrations, we manually validated suspected cross-registrations based on the primary registration, and for trials with published results, in the full-text as well as PubMed metadata and abstract. A detailed protocol of these cross-registration checks is available at https://osf.io/mjpzh.

The report cards also called for action on open access (OA) and thus required an accurate OA status of publications. The OA status of publications was determined by querying the Unpaywall API using UnpaywallR. Publications were considered as OA if the peer-reviewed version was openly accessible (without fees or log-in) on either the journal website or a repository. Social networking platforms for researchers (e.g., ResearchGate) or personal websites were not considered as longterm openly accessible venues. Unpaywall has been reported to provide a conservative estimate of the actual percentage of OA in the literature (Piwowar et al., 2018), and changes in a publication’s OA status are reflected in Unpaywall with a delay. Therefore, we performed manual checks throughout the study of the OA status of publications that were marked as closed in Unpaywall. Manual checks of the OA status of publications were limited to the following study time points: launch, 3 months post-launch, and 12 months post-launch. Any corrections in OA status based on the manual check performed at launch were also applied to the pre-intervention study point for a given publication. Moreover, any new publications shared by investigators were manually checked before sending updated report cards; in these cases, this OA status was also applied to previous study time points. A detailed protocol of these OA checks is available at https://osf.io/3upgy.

## 6 Reporting checklists

### CROSS Checklist for Reporting Of Survey Studies

Sharma, A., Minh Duc, N. T., Luu Lam Thang, T., Nam, N. H., Ng, S. J., Abbas, K. S., Huy, N. T., Marušić, A., Paul, C. L., Kwok, J., Karbwang, J., de Waure, C., Drummond, F. J., Kizawa, Y., Taal, E., Vermeulen, J., Lee, G. H. M., Gyedu, A., To, K. G., … Karamouzian, M. (2021). A Consensus-Based Checklist for Reporting of Survey Studies (CROSS). Journal of General Internal Medicine, 36(10), 3179–3187. https://doi.org/10.1007/s11606-021-06737-1

| Section/topic | Item | Item description | Reported in section [name] |
| --- | --- | --- | --- |
| **Title and abstract** |  |  |  |
| Title and abstract | 1a | State the word “survey” along with a commonly used term in title or abstract to introduce the study’s design. | Abstract |
|  | 1b | Provide an informative summary in the abstract, covering background, objectives, methods, findings/results, interpretation/discussion, and conclusions. | Abstract |
| **Introduction** |  |  |  |
| Background | 2 | Provide a background about the rationale of study, what has been previously done, and why this survey is needed. | Introduction, paragraphs 1-4 |
| Purpose/aim | 3 | Identify specific purposes, aims, goals, or objectives of the study. | Introduction, paragraphs 5-6 |
| **Methods** |  |  |  |
| Study design | 4 | Specify the study design in the methods section with a commonly used term (e.g., cross-sectional or longitudinal). | Methods/Feasibility study |
|  | 5a | Describe the questionnaire (e.g., number of sections, number of questions, number and names of instruments used). | Methods/Feasibility study/Materials/Questionnaire development and pretesting;<br>OSF project (https://osf.io/stnp5/) |
| Data collection methods | 5b | Describe all questionnaire instruments that were used in the survey to measure particular concepts. Report target population, reported validity and reliability information, scoring/classification procedure, and reference links (if any). | NA |
|  | 5c | Provide information on pretesting of the questionnaire, if performed (in the article or in an online supplement). Report the method of pretesting, number of times questionnaire was pre-tested, number and demographics of participants used for pretesting, and the level of similarity of demographics between pre-testing participants and sample population. | Supplement S1;<br>Methods/Feasibility study/Materials/Questionnaire development and pretesting;<br>OSF project (https://osf.io/stnp5/) |
|  | 5d | Questionnaire if possible, should be fully provided (in the article, or as appendices or as an online supplement). | OSF project (https://osf.io/stnp5/) |
| Sample characteristics | 6a | Describe the study population (i.e., background, locations, eligibility criteria for participant inclusion in survey, exclusion criteria). | Methods/Feasibility study/Sample development;<br>Supplement S2 |
|  | 6b | Describe the sampling techniques used (e.g., single stage or multistage sampling, simple random sampling, stratified sampling, cluster sampling, convenience sampling). Specify the locations of sample participants whenever clustered sampling was applied. | Methods/Feasibility study/Sample development |
|  | 6c | Provide information on sample size, along with details of sample size calculation. | Methods/Feasibility study/Sample development |
|  | 6d | Describe how representative the sample is of the study population (or target population if possible), particularly for population-based surveys. | Methods/Feasibility study/Sample development |
| Survey administration | 7a | Provide information on modes of questionnaire administration, including the type and number of contacts, the location where the survey was conducted (e.g., outpatient room or by use of online tools, such as SurveyMonkey). | Methods/Feasibility study/Materials/Administration of intervention and survey;<br>Methods/Feasibility study/Materials/Questionnaire development and pretesting;<br>Methods/Feasibility study/Sample development/Trial investigators<br>Supplement S3 |
|  | 7b | Provide information of survey's time frame, such as periods of recruitment, exposure, and follow-up days. | Methods/Feasibility study/Materials/Administration of intervention and survey |
|  | 7c | Provide information on the entry process:<br>—>For non-web-based surveys, provide approaches to minimize human error in data entry.<br>—>For web-based surveys, provide approaches to prevent “multiple participation” of participants. | Methods/Feasibility study/Analysis/Analysis of perceptions of the report card and infosheet |
| Study preparation | 8 | Describe any preparation process before conducting the survey (e.g., interviewers' training process, advertising the survey). | Supplement S1 |
| Ethical considerations | 9a | Provide information on ethical approval for the survey if obtained, including informed consent, institutional review board [IRB] approval, Helsinki declaration, and good clinical practice [GCP] declaration (as appropriate). | Methods/Feasibility study/Ethical approval and study protocol |
|  | 9b | Provide information about survey anonymity and confidentiality and describe what mechanisms were used to protect unauthorized access. | Methods/Feasibility study/Materials/Questionnaire development and pretesting;<br>Methods/Feasibility study/Materials/Administration of intervention and survey<br>Methods/Feasibility study/Analysis/Analysis of perceptions of the report card and infosheet |
| Statistical analysis | 10a | Describe statistical methods and analytical approach. Report the statistical software that was used for data analysis. | Methods/Feasibility study/Analysis/Analysis of perceptions of the report card and infosheet |
|  | 10b | Report any modification of variables used in the analysis, along with reference (if available). | Protocol deviations (see OSF project: https://osf.io/stnp5/) |
|  | 10c | Report details about how missing data was handled. Include rate of missing items, missing data mechanism (i.e., missing completely at random [MCAR], missing at random [MAR] or missing not at random [MNAR]) and methods used to deal with missing data (e.g., multiple imputation). | Methods/Feasibility study/Analysis/Analysis of perceptions of the report card and infosheet |
|  | 10d | State how non-response error was addressed. | Methods/Feasibility study/Analysis/Analysis of perceptions of the report card and infosheet |
|  | 10e | For longitudinal surveys, state how loss to follow-up was addressed. | NA |
|  | 10f | Indicate whether any methods such as weighting of items or propensity scores have been used to adjust for non-representativeness of the sample. | NA |
|  | 10g | Describe any sensitivity analysis conducted. | Methods/Feasibility study/Analysis/Analysis of perceptions of the report card and infosheet |
| **Results** |  |  |  |
| Respondent characteristics | 11a | Report numbers of individuals at each stage of the study. Consider using a flow diagram, if possible. | Figure 2 |
|  | 11b | Provide reasons for non-participation at each stage, if possible. | Figure 2<br>Results/Feasibility study/Sample characteristics<br>Results/Feasibility study/Survey |
|  | 11c | Report response rate, present the definition of response rate or the formula used to calculate response rate. | Results/Feasibility study/Survey;<br>Figure 2;<br>Supplement S3 |
|  | 11d | Provide information to define how unique visitors are determined. Report number of unique visitors along with relevant proportions (e.g., view proportion, participation proportion, completion proportion). | Methods/Feasibility study/Analysis/Analysis of perceptions of the report card and infosheet |
| Descriptive results | 12 | Provide characteristics of study participants, as well as information on potential confounders and assessed outcomes. | Results/Feasibility study/Sample characteristics;<br>Results/Feasibility study/Survey;<br>Supplements S2-S3 |
| Main findings | 13a | Give unadjusted estimates and, if applicable, confounder-adjusted estimates along with 95% confidence intervals and p-values. | Figure 3;<br>Supplement S3;<br>Results/Feasibility study/Survey |
|  | 13b | For multivariable analysis, provide information on the model building process, model fit statistics, and model assumptions (as appropriate). | NA |
|  | 13c | Provide details about any sensitivity analysis performed. If there are considerable amount of missing data, report sensitivity analyses comparing the results of complete cases with that of the imputed dataset (if possible). | Results/Feasibility study/Survey;<br>Supplement S3 |
| **Discussion** |  |  |  |
| Limitations | 14 | Discuss the limitations of the study, considering sources of potential biases and imprecisions, such as non-representativeness of sample, study design, important uncontrolled confounders. | Discussion/Strengths, limitations, and challenges |
| Interpretations | 15 | Give a cautious overall interpretation of results, based on potential biases and imprecisions and suggest areas for future research. | Discussion/Summary of findings/Perceptions of the report card and infosheet;<br>Discussion/Implications and future work |
| Generalizability | 16 | Discuss the external validity of the results. | Discussion/Research in context |
| **Other sections** |  |  |  |
| Role of funding source | 17 | State whether any funding organization has had any roles in the survey's design, implementation, and analysis. | Funding |
| Conflict of interest | 18 | Declare any potential conflict of interest. | Declarations |
| Acknowledgements | 19 | Provide names of organizations/persons that are acknowledged along with their contribution to the research. | Acknowledgements |

### CONSORT Checklist for Pilot and Feasibility Trials

Eldridge, S. M., Chan, C. L., Campbell, M. J., Bond, C. M., Hopewell, S., Thabane, L., Lancaster, G. A., Altman, D., Bretz, F., Campbell, M., Cobo, E., Craig, P., Davidson, P., Groves, T., Gumedze, F., Hewison, J., Hirst, A., Hoddinott, P., Lamb, S. E., … on behalf of the PAFS consensus group. (2016). CONSORT 2010 statement: Extension to randomised pilot and feasibility trials. Pilot and Feasibility Studies, 2(1), 64. https://doi.org/10.1186/s40814-016-0105-8.

Table adapted from: https://figshare.com/articles/journal_contribution/CONSORT_2010_checklist_of_information_to_include_when_reporting_a_pilot_or_feasibility_trial_/21519771.

| Section/Topic | Item No | Checklist item | Reported in section [name] |
| --- | --- | --- | --- |
| **Title and abstract** |  |  |  |
|  | 1a | Identification as a pilot or feasibility randomised trial in the title | Title |
|  | 1b | Structured summary of pilot trial design, methods, results, and conclusions (for specific guidance see CONSORT abstract extension for pilot trials) | Abstract |
| **Introduction** |  |  |  |
| Background and objectives | 2a | Scientific background and explanation of rationale for future definitive trial, and reasons for randomised pilot trial | Introduction, paragraphs 1-4 |
|  | 2b | Specific objectives or research questions for pilot trial | Introduction, paragraphs 5-6 |
| **Methods** |  |  |  |
| Trial design | 3a | Description of pilot trial design (such as parallel, factorial) including allocation ratio | Methods/Feasibility study |
|  | 3b | Important changes to methods after pilot trial commencement (such as eligibility criteria), with reasons | Protocol deviations (see OSF project: https://osf.io/stnp5/) |
| Participants | 4a | Eligibility criteria for participants | Methods/Feasibility study/Sample development;<br>Supplement S2 |
|  | 4b | Settings and locations where the data were collected | Methods/Feasibility study/Materials/Administration of intervention and survey |
|  | 4c | How participants were identified and consented | Methods/Feasibility study/Sample development/Trial investigators;<br>Methods/Feasibility study/Materials/Administration of intervention and survey<br>OSF project: https://osf.io/stnp5/ |
| Interventions | 5 | The interventions for each group with sufficient details to allow replication, including how and when they were actually administered | Methods/Feasibility study/Materials/Administration of intervention and survey; |
|  |  |  | Methods/Tools: report cards and infosheet;<br>OSF project:<br>https://osf.io/stnp5/ |
| Outcomes | 6a | Completely defined prespecified assessments or measurements to address each pilot trial objective specified in 2b, including how and when they were assessed | Methods/Feasibility study/Analysis/Analysis of improvement in transparency practices<br>Preregistration in OSF:<br>https://osf.io/stnp5/ |
|  | 6b | Any changes to pilot trial assessments or measurements after the pilot trial commenced, with reasons | Protocol deviations (see OSF project:<br>https://osf.io/stnp5/) |
|  | 6c | If applicable, prespecified criteria used to judge whether, or how, to proceed with future definitive trial | NA |
| Sample size | 7a | Rationale for numbers in the pilot trial | Methods/Feasibility study;<br>Methods/Feasibility study/Sample development |
|  | 7b | When applicable, explanation of any interim analyses and stopping guidelines | NA |
| Randomisation: |  |  |  |
| Sequence generation | 8a | Method used to generate the random allocation sequence | NA |
|  | 8b | Type of randomisation(s); details of any restriction (such as blocking and block size) | NA |
| Allocation Concealment mechanism | 9 | Mechanism used to implement the random allocation sequence (such as sequentially numbered containers), describing any steps taken to conceal the sequence until interventions were assigned | NA |
| Implementation | 10 | Who generated the random allocation sequence, who enrolled participants, and who assigned participants to interventions | NA |
| Blinding | 11a | If done, who was blinded after assignment to interventions (for example, participants, care providers, those assessing outcomes) and how | NA |
|  | 11b | If relevant, description of the similarity of interventions | NA |
| Statistical methods | 12 | Methods used to address each pilot trial objective whether qualitative or quantitative | Methods/Feasibility study/Analysis/Analysis of improvement in transparency practices |
| **Results** |  |  |  |
| Participant flow (a diagram is strongly recommended) | 13a | For each group, the numbers of participants who were approached and/or assessed for eligibility, randomly assigned, received intended treatment, and were assessed for each objective | Figure 2 |
|  | 13b | For each group, losses and exclusions after randomisation, together with reasons | Figure 2;<br>Results/Feasibility study/Sample characteristics |
| Recruitment | 14a | Dates defining the periods of recruitment and follow-up | Supplement S3 |
|  | 14b | Why the pilot trial ended or was stopped | NA |
| Baseline data | 15 | A table showing baseline demographic and clinical characteristics for each group | Supplement S2 |
| Numbers analysed | 16 | For each objective, number of participants (denominator) included in each analysis. If relevant, these numbers should be by randomised group | Figure 2 |
| Outcomes and estimation | 17 | For each objective, results including expressions of uncertainty (such as 95% confidence interval) for any estimates. If relevant, these results should be by randomised group | Figure 4;<br>Results/Feasibility study/Improvement in transparency practices |
| Ancillary analyses | 18 | Results of any other analyses performed that could be used to inform the future definitive trial | Supplement S3 |
| Harms | 19 | All important harms or unintended effects in each group (for specific guidance see CONSORT for harms) | NA |
|  | 19a | If relevant, other important unintended consequences | NA |
| **Discussion** |  |  |  |
| Limitations | 20 | Pilot trial limitations, addressing sources of potential bias and remaining uncertainty about feasibility | Discussion/Strengths, limitations, and challenges |
| Generalisability | 21 | Generalisability (applicability) of pilot trial methods and findings to future definitive trial and other studies | Discussion/Scaling beyond transparency in clinical trials;<br>Discussion/Implications and future work |
| Interpretation | 22 | Interpretation consistent with pilot trial objectives and findings, balancing potential benefits and harms, and considering other relevant evidence | Discussion/Summary of findings/Transparency improvements in practice;<br>Discussion/Research in context |
|  | 22a | Implications for progression from pilot to future definitive trial, including any proposed amendments | Discussion/Implications and future work |
| **Other information** |  |  |  |
| Registration | 23 | Registration number for pilot trial and name of trial registry | NA |
| Protocol | 24 | Where the pilot trial protocol can be accessed, if available | Methods/Feasibility study/Ethical approval and study protocol |
| Funding | 25 | Sources of funding and other support (such as supply of drugs), role of funders | Funding |
|  | 26 | Ethical approval or approval by research review committee, confirmed with reference number | Methods/Feasibility study/Ethical approval and study protocol |

